# Effect of Low-Dose Rivaroxaban Plus Aspirin on Carotid Plaque Inflammation (SPIRIT): A Randomized ^18^F-FDG PET/CT Trial

**DOI:** 10.1101/2025.11.18.25340536

**Authors:** Tae Oh Kim, Sangwon Han, Jung Bok Lee, Chong Hyun Suh, Soo Jin Kang, Dae Hyuk Moon, Cheol Whan Lee, Jun Gyo Gwon, Seung-Whan Lee, the SPIRIT Investigators

**Author notes:** The first two authors (Drs. TO. Kim and S. Han) contributed equally to this article. Corresponding Author: **Address for correspondence**: Dr. Seung-Whan Lee, Division of Cardiolgy, Department of Internal Medicine, Asan Medical Center, University of Ulsan College of Medicine, 88, Olympic-ro 43-gil, Songpa-gu, Seoul 05505, Korea., Dr. Jun Gyo Gwon, Division of Vascular Surgery, Department of Surgery, Asan Medical Center, University of Ulsan College of Medicine, 88, Olympic-ro 43-gil, Songpa-gu, Seoul 05505, Korea.

## Abstract

**Background:** Aspirin plus rivaroxaban reduces cardiovascular events in patients with stable atherosclerosis; however, whether Factor Xa inhibition attenuates arterial wall inflammation in humans remains unclear.

**Methods:** We conducted a single-center, randomized, open-label, proof-of-concept study with blinded endpoint assessment using ¹⁸F-fluorodeoxyglucose positron emission tomography/computed tomography (¹⁸F-FDG PET/CT). Adults with asymptomatic carotid stenosis (20–80%) and baseline carotid inflammation (target-to-background ratio [TBR] ≥1.6) were randomly assigned in a 1:1 ratio to receive either aspirin (100 mg/day) plus rivaroxaban (2.5 mg twice daily) or aspirin (100 mg/day) alone. The primary endpoint was the percent change in the most diseased segment (MDS) TBR of the index carotid artery from baseline to 12 months. Secondary endpoints included whole-vessel carotid TBR, aortic TBR, and changes in lipids and high-sensitivity C-reactive protein.

**Results:** Between September 2021 and December 2023, 92 patients were randomized (mean age 69.4 years; 88% men); 81 (88%) completed 12-month imaging. Mean percent changes in carotid MDS TBR were −7.25% (95% CI, −11.03 to −3.48) and −7.36% (95% CI, −11.06 to −3.67) in the combination therapy and aspirin-alone groups, respectively. The adjusted between-group difference (combination therapy minus aspirin alone) was −1.50 percentage points (95% CI, −6.20 to 3.21; P=0.53). No significant between-group differences were observed in whole-vessel carotid or aortic TBR, or in laboratory parameters. Minor bleeding occurred in four patients (8.7%) receiving combination therapy and in one (2.2%) receiving aspirin alone, with no major bleeding or deaths.

**Conclusions:** In patients with carotid atherosclerosis, adding low-dose rivaroxaban to aspirin did not reduce arterial inflammation beyond aspirin alone over 12 months on ¹⁸F-FDG PET/CT.

**Clinical Trial Registration:** ClinicalTrials.gov NCT05797376

**CLINICAL PERSPECTIVE:** *What is new?:*

- This is the first randomized trial using serial ^18^F-FDG PET/CT imaging to directly evaluate whether dual-pathway inhibition with low-dose rivaroxaban plus aspirin reduces arterial inflammation compared with aspirin alone.
- Despite proven cardiovascular benefits in the COMPASS trial, the addition of rivaroxaban to aspirin did not confer additional anti-inflammatory effects on human atherosclerotic plaques over 12 months.
- Both treatment groups showed similar reductions in arterial inflammation, likely reflecting the anti-inflammatory effects of background statin therapy and aspirin.

*What are the clinical implications?:*

- The cardiovascular benefits of dual-pathway inhibition are likely mediated primarily by prevention of thrombotic events rather than by plaque stabilization through reduced inflammation.
- Optimal cardiovascular risk reduction requires a complementary approach: antithrombotic therapy to prevent acute events and anti-inflammatory interventions to stabilize atherosclerotic plaques.
- Future studies should explore whether combining dual-pathway inhibition with targeted anti-inflammatory therapies (such as colchicine, IL-1β inhibition) provides synergistic cardiovascular protection.

## INTRODUCTION

Atherosclerotic cardiovascular disease remains the leading cause of death globally.^1,2^ Acute events result from rupture or erosion of vulnerable, inflammation-rich plaques followed by thrombosis.^3^ This pathology supports therapeutic strategies targeting both inflammation and thrombosis.^4,5^

Dual-pathway inhibition with low-dose rivaroxaban (2.5 mg twice daily) plus aspirin reduces major adverse cardiovascular events compared with aspirin alone in patients with stable atherosclerotic disease.^6,7^ Beyond its antithrombotic effects, Factor Xa can activate protease-activated receptor (PAR)–dependent inflammatory signaling independent of thrombin generation, suggesting that Factor Xa inhibition could attenuate plaque inflammation.^8^ Whether rivaroxaban exerts anti-inflammatory effects within human atherosclerosis remains unknown.

¹⁸F-fluorodeoxyglucose positron emission tomography/computed tomography (¹⁸F-FDG PET/CT) enables quantitative assessment of arterial wall inflammation by indexing macrophage glucose metabolism. Arterial FDG uptake correlates with histologic inflammation, predicts cardiovascular events, and is widely used as an intermediate imaging endpoint for evaluating anti-inflammatory therapies.^9,10^

We conducted the SPIRIT (Effects of aspirin versus aspirin plus low-dose rivaroxaban on carotid atherosclerotic plaque inflammation) trial to test the hypothesis that rivaroxaban 2.5 mg twice daily plus aspirin would reduce carotid plaque inflammation compared with aspirin alone in patients with stable carotid atherosclerosis, as assessed by serial ¹⁸F-FDG PET/CT.

## METHODS

The data that support the findings of this study are available from the corresponding authors on reasonable request.

### Trial Design and Oversight

SPIRIT was an investigator-initiated, prospective, randomized, open-label trial with blinded endpoint assessment (PROBE design) conducted at Asan Medical Center (Seoul, Republic of Korea) between September 2021 and December 2023. This proof-of-concept mechanistic study evaluated the anti-inflammatory effects of rivaroxaban plus aspirin using quantitative vascular imaging. The study protocol was approved by the institutional review board of Asan Medical Center, and the trial complied with the Declaration of Helsinki and Good Clinical Practice guidelines. All participants provided written informed consent prior to enrollment. The trial was registered at ClinicalTrials.gov (NCT05797376).

The study was funded by Bayer Korea, which provided the study medication and financial support but had no role in study design, data collection, analysis, interpretation, or manuscript preparation. An independent data and safety monitoring board reviewed unblinded safety data, and an independent clinical events committee adjudicated outcomes while remaining blinded to treatment assignment. Data management and statistical analyses were performed by the Clinical Research Center of Cardiology at Asan Medical Center. All authors had full access to the data and approved the final manuscript. Participating investigators are listed in the Supplementary Material (Section A), and details regarding trial organization and oversight are provided in Sections B and C.

### Study Population and Randomization

Eligible participants were adults (≥18 years) with asymptomatic carotid artery stenosis (20–80% diameter stenosis by duplex ultrasound) and established atherosclerotic cardiovascular disease meeting the Cardiovascular Outcomes for People Using Anticoagulation Strategies (COMPASS) trial eligibility criteria.^6^ To ensure adequate baseline inflammation, only patients with screening ¹⁸F-FDG PET/CT evidence of carotid inflammation—defined as a most diseased segment (MDS) target-to-background ratio (TBR) ≥ 1.6—were randomized.^11^ Full inclusion and exclusion criteria are provided in the Supplementary Material (Section D)

Participants were randomly assigned in a 1:1 ratio to receive either rivaroxaban (2.5 mg twice daily) plus aspirin (100 mg/day) or aspirin (100 mg/day) alone. Randomization was performed using a secure web-based system with permuted blocks of four. Allocation was concealed and released only after eligibility confirmation and baseline data entry.

### ^18^F-FDG PET/CT Imaging and Analysis

All patients underwent ¹⁸F-FDG PET/CT imaging at baseline and 12 months. Patients fasted for ≥8 h, and blood glucose was confirmed to be <150 mg/dL before tracer administration. ¹⁸F-FDG was administered intravenously at 5.2 MBq (0.14 mCi) per kilogram. Three-dimensional PET/CT was performed 2 h after injection (standard uptake time) using one of three scanners — Discovery 690, Discovery 690 Elite, or Discovery 710 (GE Healthcare, Waukesha, WI). Standardized uptake values (SUVs) were harmonized across scanners through routine calibration using a dose calibrator (CRC-25 PET; Capintec Inc., Florham Park, NJ), and quality control, including daily constancy, quarterly linearity, annual accuracy and precision checks. Cross-calibration between scanners was performed annually with the same dose calibrator, and quarterly assessments of recovery coefficients were carried out for all hot cylinders of an American College of Radiology (ACR)–accredited PET phantom (Flangeless Esser PET phantom; Biodex Medical Systems Inc., Shirley, NY) without additional smoothing. A noncontrast CT scan was obtained for attenuation correction and anatomic localization, followed by a 10-min PET acquisition encompassing the carotid arteries and ascending aorta.^12^

All image analyses were performed by a single experienced nuclear medicine physician blinded to treatment assignment and clinical data. Vascular FDG uptake was quantified in both carotid arteries and the ascending aorta (from the aortic valve to the brachiocephalic trunk). Two-dimensional regions of interest were manually delineated on co-registered CT images and transferred to PET images. Segment-level TBR was calculated as arterial wall SUVmax divided by venous blood-pool SUVmean (superior vena cava).^13,14^ Whole-vessel TBR was defined as the mean of all segment TBRs, whereas MDS TBR was the mean of five consecutive segment TBRs centered on the highest-uptake region. Detailed acquisition, reconstruction, and analysis procedures are provided in the Supplementary Material (Section E) and Figure S1.

### Study Endpoints and Follow-Up

The primary endpoint was the percent change in the index carotid artery MDS TBR from baseline to 12 months. The index carotid artery was defined as the vessel with the highest baseline FDG uptake. Secondary imaging endpoints included percent changes in index carotid whole-vessel TBR and ascending aorta MDS and whole-vessel TBR. Secondary laboratory endpoints included changes in high-sensitivity C-reactive protein (hsCRP), total cholesterol, low-density lipoprotein (LDL) cholesterol, high-density lipoprotein (HDL) cholesterol, and triglycerides.

Safety endpoints included death, intracranial hemorrhage, and bleeding events classified based on the Bleeding Academic Research Consortium (BARC) criteria: major (BARC 3–5) and minor (BARC 1–2) bleeding.^15,16^ All clinical events were adjudicated by an independent committee blinded to treatment assignment. Detailed endpoint definitions are provided in the Supplementary Material (Sections F and G).

Follow-up visits were scheduled at 1, 6, and 12 months to assess adverse events, concomitant medications, and treatment adherence. Adherence was predefined as ≥80% medication intake by pill count; patients with <80% adherence were excluded from per-protocol analyses. Laboratory assessments and repeat ¹⁸F-FDG PET/CT were performed at 12 months.

### Statistical Analysis

Sample size was calculated to detect a 5-percentage point between-group difference in percent change of TBR, based on prior ¹⁸F-FDG PET/CT studies of anti-inflammatory interventions,^17^ assuming a standard deviation (SD) of 8%, two-sided α=0.05, and 80% power. This yielded 41 patients per group; accounting for approximately 10% attrition, we targeted 46 per group (N=92). Details are provided in the Supplementary Material (Section H).

The primary analysis included all randomized patients with evaluable baseline and 12-month ¹⁸F-FDG PET/CT data (modified intention-to-treat). Per-protocol sensitivity analyses included patients with ≥80% treatment adherence. Safety analyses encompassed all randomized patients.

The primary endpoint was the percent change in MDS TBR from baseline to 12 months, defined as Δ% = 100 × (TBR_12mo_-TBR_baseline_) / TBR_baseline_. Continuous variables are presented as mean±SD and compared using independent t-tests or Wilcoxon rank-sum tests, as appropriate. Categorical variables are presented as numbers (percentages) and compared using χ² or Fisher’s exact tests. Within-group changes were assessed using paired t-tests.

Between-group differences were evaluated in two ways: (1) an unadjusted Welch’s t-test comparing mean Δ%; and (2) an ANCOVA of Δ% adjusted for baseline TBR and treatment as covariates to improve precision. Secondary imaging endpoints (whole-vessel TBR of the index carotid artery; MDS and whole-vessel TBR of the ascending aorta) and laboratory parameters (lipid profile, hsCRP) were analyzed using the same framework. Prespecified subgroup analyses examined categorical effect modifiers (age, sex, diabetes, hypertension, previous stroke, ever smoker, high-intensity statin use, hsCRP, HDL, and LDL) using treatment-by-subgroup interaction terms. Exploratory, continuous-scale effect-modification analyses modeled baseline TBR with restricted cubic splines (df=4) and tested the joint significance of treatment×spline terms by a Wald F-test. No imputation was performed for missing data. Additional statistical details are provided in the Supplementary Material (Section I).

All tests were two-sided with α=0.05. No adjustment for multiplicity was applied; therefore, secondary endpoints were considered exploratory. Statistical analyses were performed using Python version 3.10 (Python Software Foundation) and R version 4.0 (R Foundation for Statistical Computing).

## RESULTS

### Study Population and Baseline Characteristics

Between September 2021 and December 2023, 133 patients were screened; 41 were excluded, and 92 were randomized in a 1:1 ratio to receive either rivaroxaban (2.5 mg twice daily) plus aspirin (100 mg/day) (n=46) or aspirin (100 mg/day) alone (n=46) (Figure 1). The primary imaging endpoint at 12 months was evaluable in 81 patients (88%): 40 in the combination-therapy group and 41 in the aspirin-alone group. The most common reason for missing imaging data was withdrawal of consent (n=8). One additional patient in the combination group did not complete imaging owing to a non-major bleeding event.

**Figure 1.**
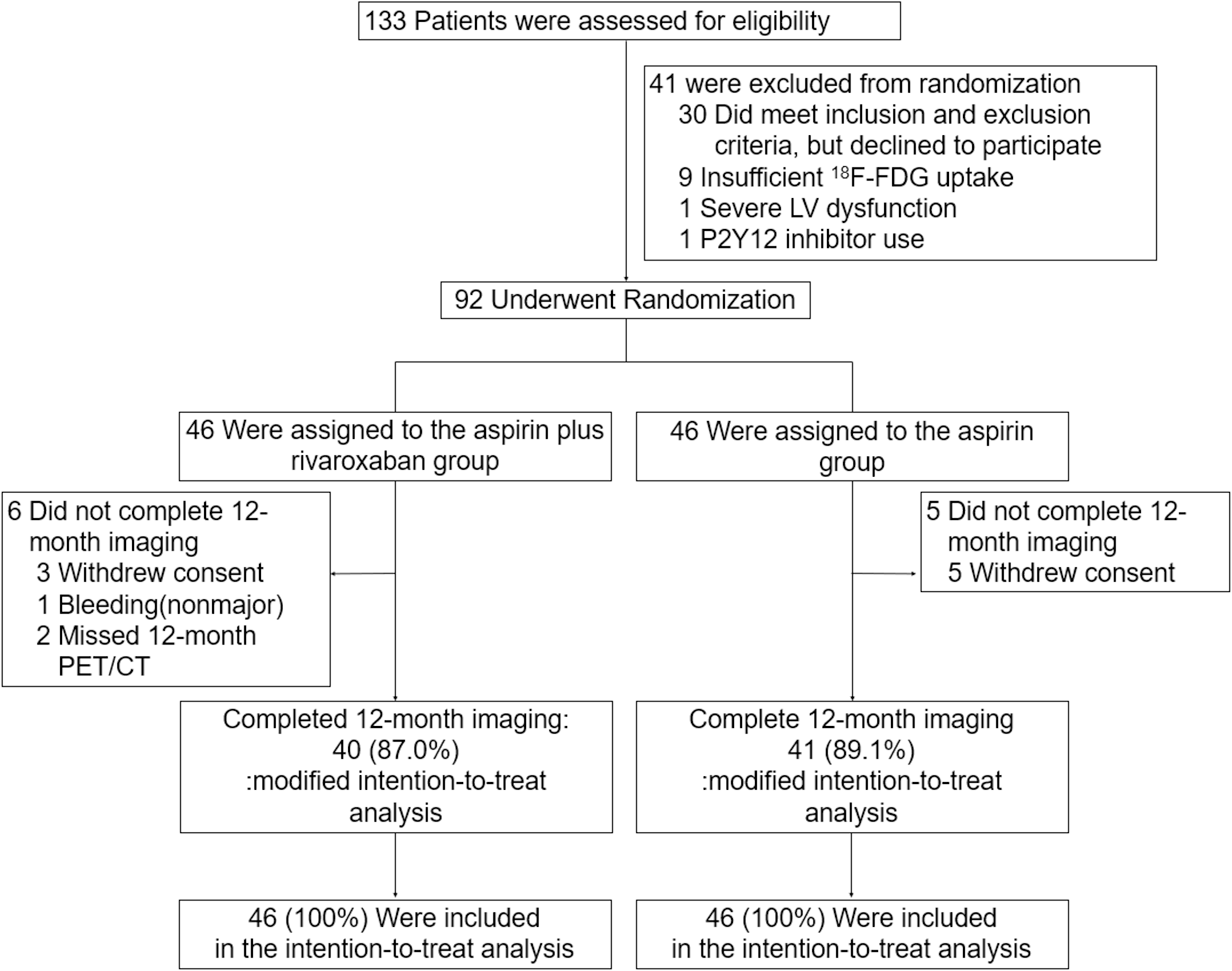
CONSORT Flow Diagram. Of 133 patients assessed for eligibility, 41 were excluded (30 met eligibility but declined participation; nine had insufficient ^18^F-FDG uptake; one had severe left ventricular dysfunction; and one was using a P2Y12 inhibitor). Ninety-two were randomized (46 to rivaroxaban plus aspirin and 46 to aspirin). Twelve-month imaging was completed in 40 (87.0%) and 41 (89.1%) patients, respectively, constituting the modified intention-to-treat (mITT) population. All 92 randomized patients were included in the intention-to-treat (ITT) analysis. **Abbreviations:** ^18^F-FDG PET/CT, fluorine-18 fluorodeoxyglucose positron emission tomography/computed tomography; MDS, most diseased segment; TBR, target-to-background ratio; ITT, intention-to-treat.

Baseline characteristics were well balanced between groups (Table 1). The mean age was 69.4±6.5 years, and 88% were men. Diabetes mellitus was present in 35.9% of patients and hypertension in 62.0%; the mean carotid stenosis was 46%. Baseline ¹⁸F-FDG PET/CT demonstrated similar vascular inflammation between groups: index carotid MDS TBR 1.99±0.41 versus 2.10±0.54, and ascending aorta MDS TBR 2.22±0.40 versus 2.32±0.42 (combination versus aspirin alone). Most participants were receiving moderate– or low-intensity statins (80.4%) and renin-angiotensin system blockers (54.4%).

**Table 1.** Baseline Characteristics.

| Characteristic | All Patients<br>(N=92) | Aspirin plus<br>Rivaroxaban<br>(N=46) | Aspirin Alone<br>(N=46) | P-value |
| --- | --- | --- | --- | --- |
| Age, y | 69.4±6.5 | 69.4±6.3 | 69.3±6.8 | 0.95 |
| Male sex, n (%) | 81 (88.0) | 41 (89.1) | 40 (87.0) | 0.75 |
| Body mass index<sup>*</sup> | 24.4±2.9 | 24.5±2.8 | 24.3±3.1 | 0.83 |
| Medical History |  |  |  |  |
| Hypertension, n (%) | 57 (62.0) | 29 (63.0) | 28 (60.9) | 0.83 |
| Diabetes mellitus, n (%) | 33 (35.9) | 16 (34.8) | 17 (37.0) | 0.83 |
| Hyperlipidemia, n (%) | 58 (63.0) | 28 (60.9) | 30 (65.2) | 0.67 |
| Current smoker, n (%) | 10 (10.9) | 5 (10.9) | 5 (10.9) | 1.00 |
| Previous stroke, n (%) | 13 (14.1) | 8 (17.4) | 5 (10.9) | 0.37 |
| Previous PCI, n (%) | 35 (38.0) | 16 (34.8) | 19 (41.3) | 0.52 |
| Previous MI, n (%) | 14 (15.2) | 6 (13.0) | 8 (17.4) | 0.56 |
| Carotid Stenosis Degree |  |  |  |  |
| Right, % | 45.9±14.2 | 48.5±15.0 | 43.2±12.9 | 0.07 |
| Left, % | 45.2±17.8 | 44.7±13.9 | 45.6±21.1 | 0.80 |
| TBR of the index vessel |  |  |  |  |
| Most diseased segment | 2.05±0.48 | 1.99±0.41 | 2.10±0.54 | 0.27 |
| Whole-vessel | 1.75±0.32 | 1.70±0.26 | 1.80±0.36 | 0.13 |
| TBR of the ascending aorta |  |  |  |  |
| Most diseased segment | 2.27±0.41 | 2.22±0.40 | 2.32±0.42 | 0.29 |
| Whole-vessel | 2.05±0.35 | 2.01±0.35 | 2.09±0.34 | 0.27 |
| Current Medication |  |  |  |  |
| RAS blocker | 50 (54.4) | 28 (60.9) | 22 (47.8) | 0.21 |
| Statin |  |  |  |  |
| High-intensity | 15 (16.3) | 7 (15.2) | 8 (17.4) | 0.84 |
| Moderate/low- intensity | 74 (80.4) | 38 (82.6) | 36 (78.3) |  |
| Ezetimibe | 60 (65.2) | 29 (63.0) | 31 (67.4) | 0.66 |

Data are presented as mean±SD or number (%). Analysis includes all 92 randomized patients.
Percentages may not sum to 100% because of rounding.
\*Body mass index is calculated as weight in kilograms divided by the square of height in meters.
Abbreviations: MI, myocardial infarction; PCI, percutaneous coronary intervention; RAS, renin-angiotensin system; TBR, target-to-background ratio.

### Primary Endpoint

At 12 months, index carotid MDS TBR decreased from 1.99±0.41 to 1.82±0.35 in the combination group and from 2.10±0.54 to 1.95±0.44 in the aspirin-alone group (Table 2). Both groups showed significant within-group reductions (P<0.01 and P<0.001, respectively), as illustrated in representative cases (Figure 2) and individual patient trajectories (Figure 3A).

**Figure 2.**
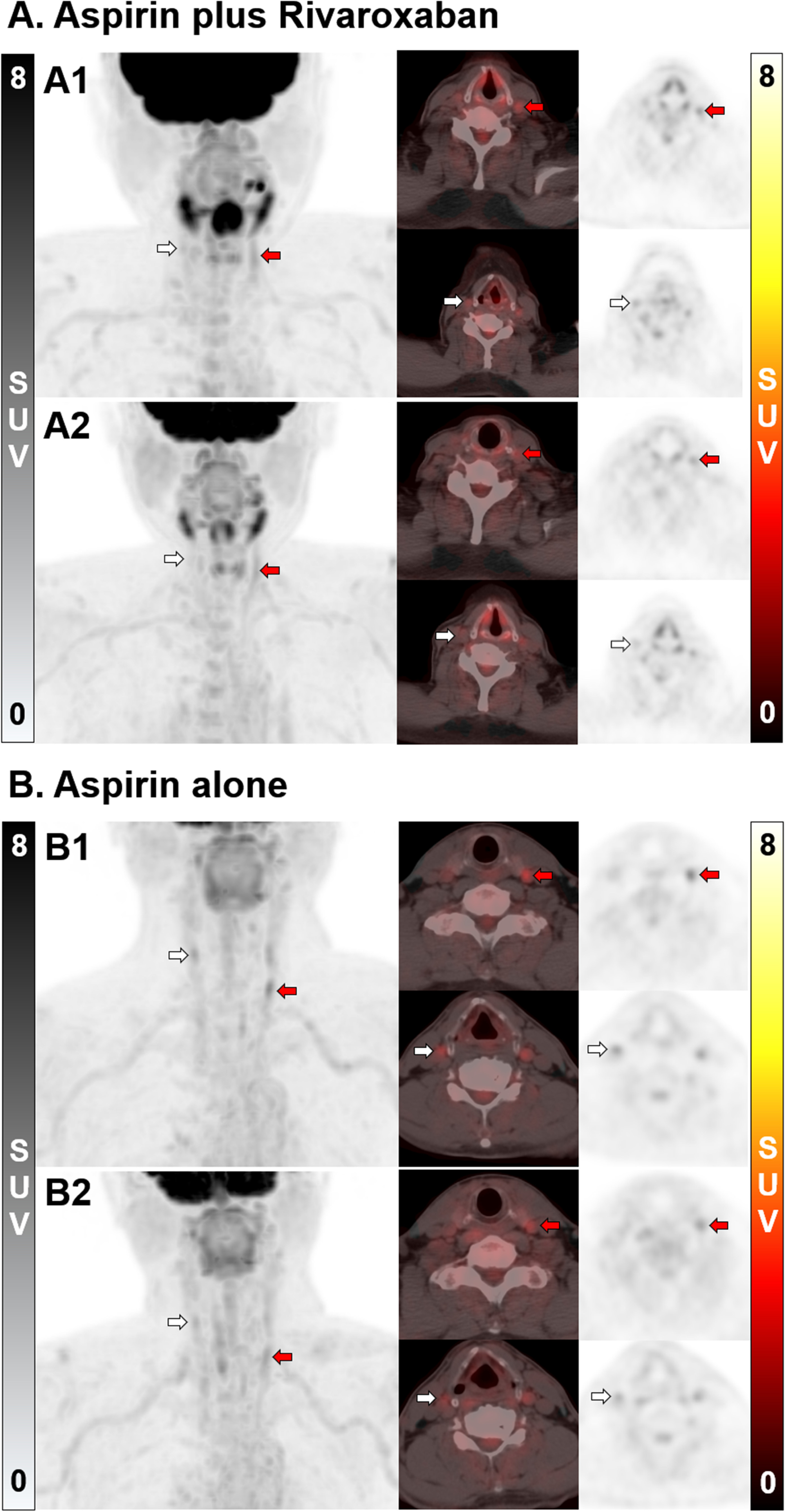
Representative Baseline and Follow-Up ^18^F-FDG PET/CT Images of the Carotid Arteries. (A) Rivaroxaban plus aspirin group: Patients A1 and A2 showing coronal PET (left), axial PET/CT fusion (middle), and axial PET (right) images. Red arrows indicate the index carotid arteries with increased FDG uptake at baseline, which decreased after treatment. White arrows indicate contralateral carotid arteries without significant uptake. (B) Aspirin alone group: Patients B1 and B2 with the same imaging sequences. SUV scale 0‒8 shown on the right. Images demonstrate a reduction in vascular inflammation (measured as MDS values) from baseline to 12-month follow-up. **Abbreviations:** ^18^F-FDG PET/CT, fluorine-18 fluorodeoxyglucose positron emission tomography/computed tomography

**Figure 3.**
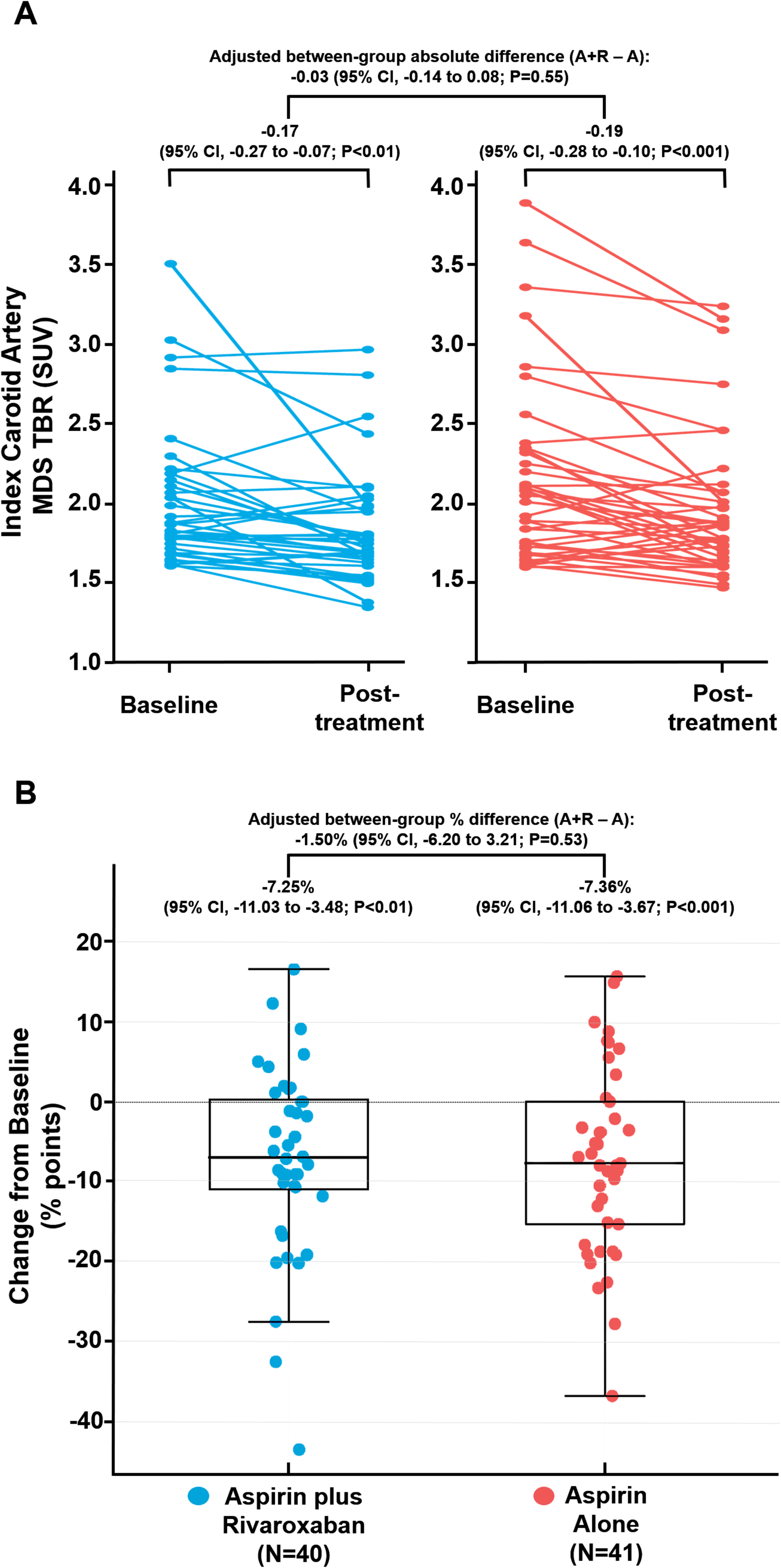
Changes in Carotid Artery Inflammation. (A) Individual patient trajectories of TBR in the MDS of the index carotid artery from baseline to 12 months. (B) Percent change from baseline in MDS TBR by treatment group. Box plots display median (line), interquartile range (box), and range (whiskers). Individual patient data are shown as colored dots. Between-group differences are adjusted for baseline TBR. Unadjusted values are provided in Table 2. **Abbreviations:** MDS, most diseased segment; TBR, target-to-background ratio

**Table 2.**
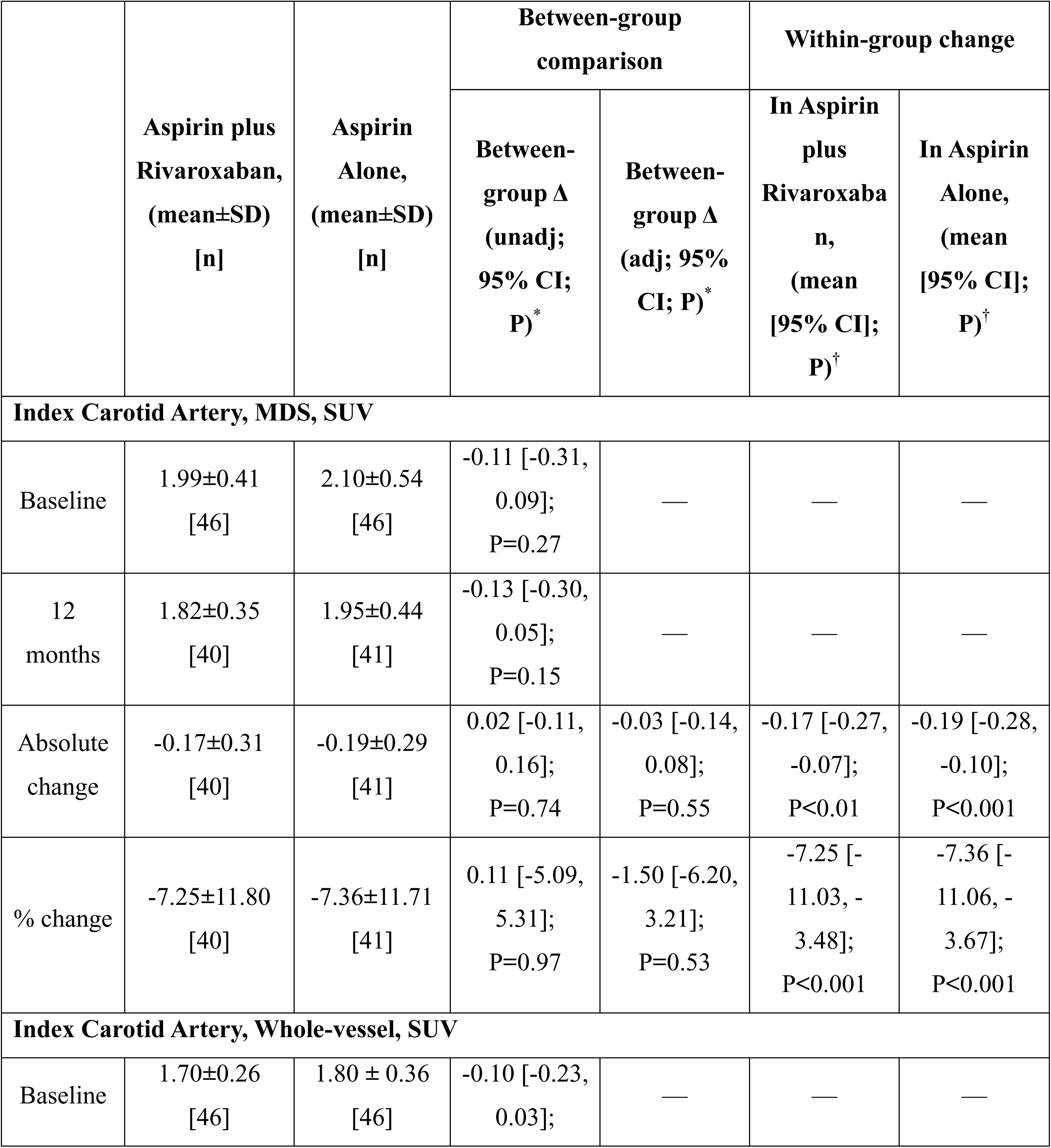
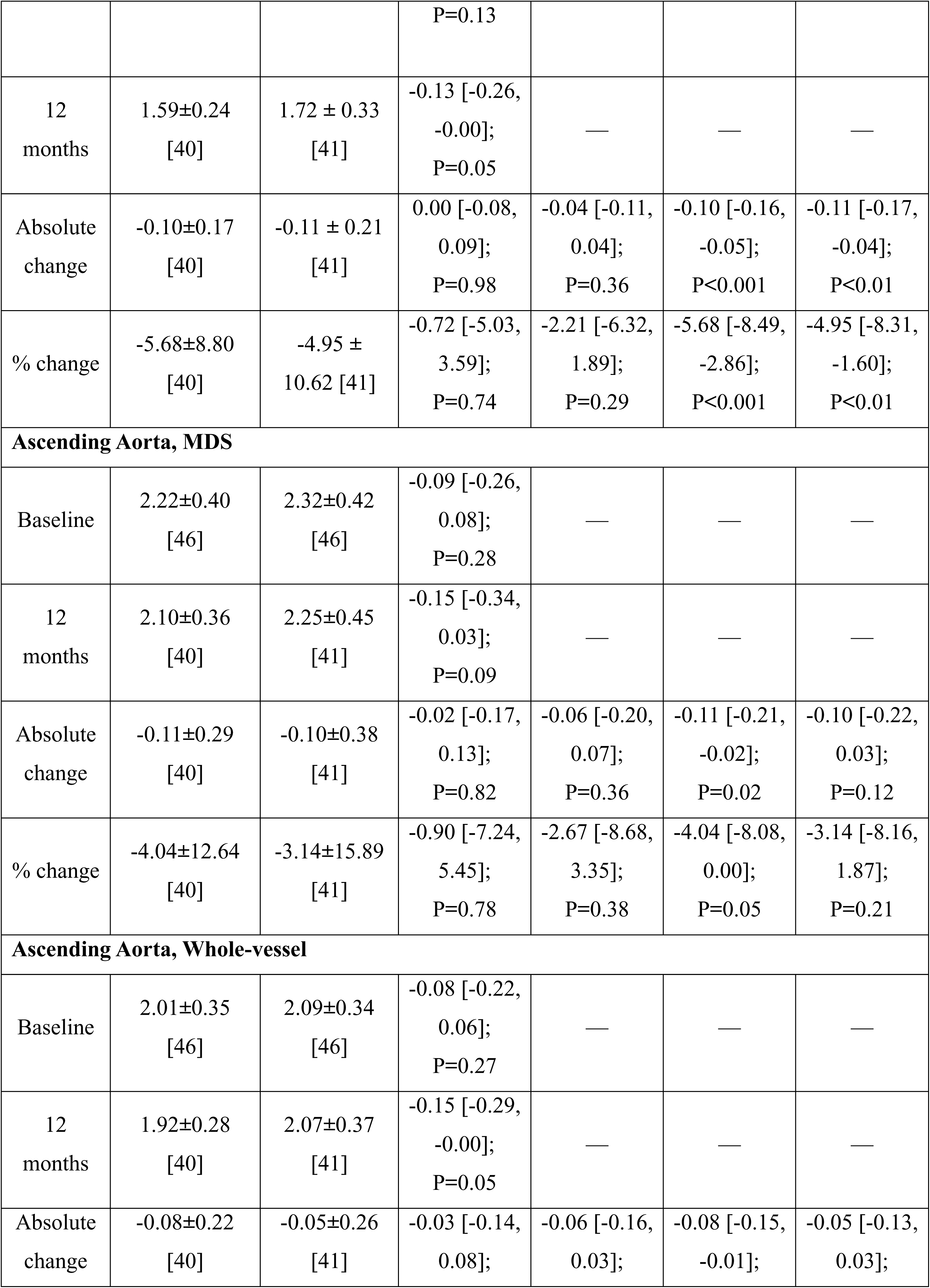
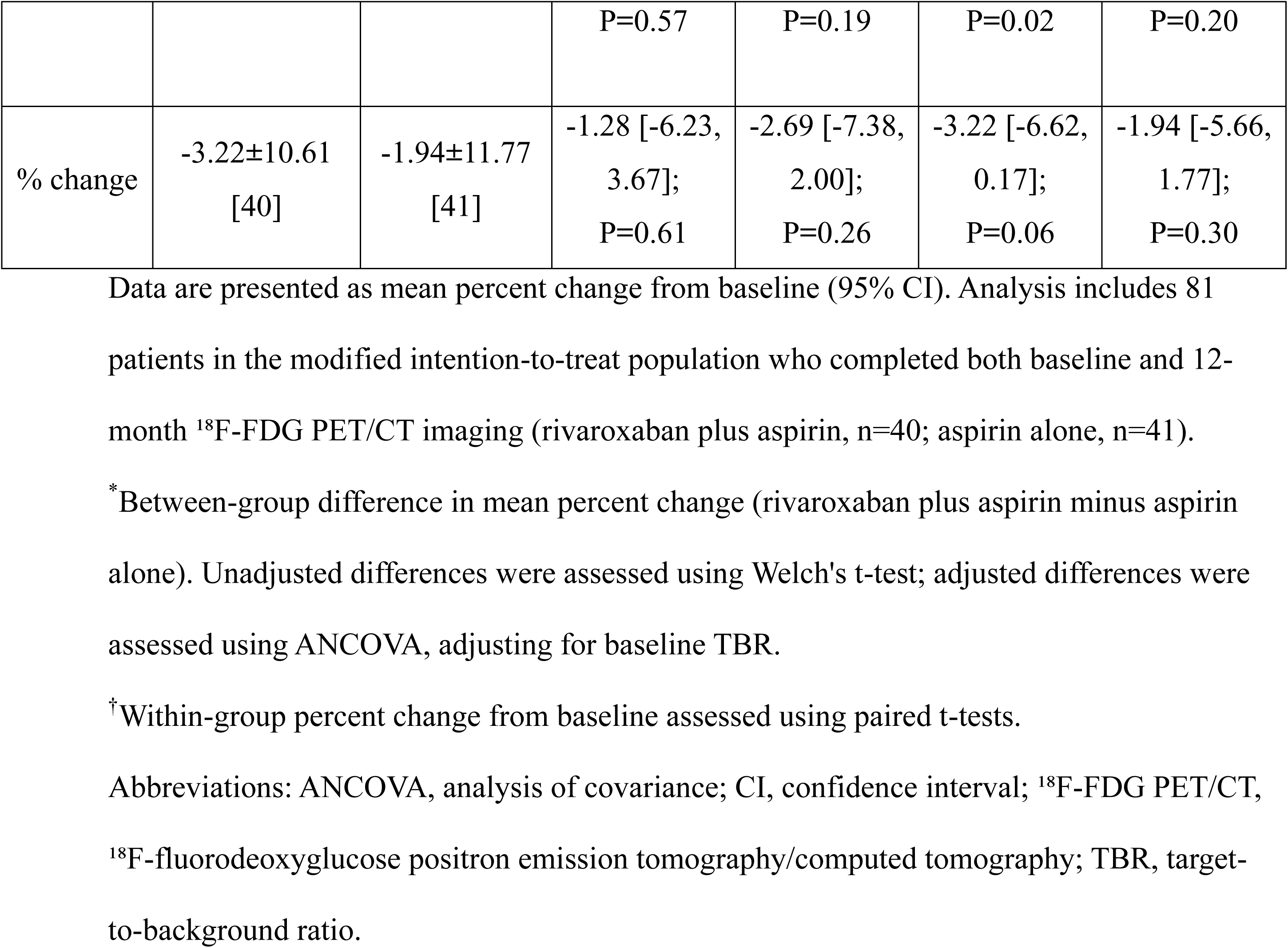
Percentage Change in TBR From Baseline to 12 Months.

However, no between-group difference was observed in percent change of MDS TBR (Table 2; Figure 3B): −7.25% (95% CI, −11.03 to −3.48) with combination therapy versus −7.36% (95% CI, −11.06 to −3.67) with aspirin alone (adjusted difference, −1.50% [95% CI, −6.20 to 3.21]; P=0.53).

### Secondary Endpoints

Secondary imaging endpoints were consistent with the primary endpoint (Table 2; Figure S2). For the index carotid, whole-vessel TBR declined by 5.68% with combination therapy and 4.95% with aspirin alone (adjusted difference, −2.21%; P=0.29). In the ascending aorta, within-group reductions were observed with combination therapy (MDS, −4.04% [P=0.02]; whole-vessel, −3.23% [P=0.02]), whereas changes with aspirin alone were not statistically significant (MDS, −3.14% [P=0.12]; whole-vessel, −1.94% [P=0.20]). Between-group differences remained nonsignificant (adjusted differences: MDS, −2.67% [P=0.38]; whole-vessel, −2.69% [P=0.26]). All secondary P values are nominal without multiplicity adjustment.

Laboratory parameters showed no between-group differences at 12 months (Table 3). HDL cholesterol increased modestly in both groups; however, between-group comparisons were non-significant. Total cholesterol, LDL cholesterol, and triglycerides did not change substantially. In contrast, hsCRP decreased slightly with combination therapy (−0.01±0.14 mg/dL) and increased with aspirin alone (0.13±0.42 mg/L), yielding an adjusted between-group difference of −0.15 mg/L (95% CI, −0.28 to −0.01; P=0.03).

**Table 3.** Changes in Laboratory Parameters from Baseline to 12 Months.

|  | Aspirin plus<br>Rivaroxaban,<br>(mean±SD)<br>[n] | Aspirin<br>Alone,<br>(mean±SD)<br>[n] | Between-group comparison |  | Within-group change |  |
| --- | --- | --- | --- | --- | --- | --- |
|  |  |  | Between-group Δ<br>(unadj; 95%<br>CI; P)* | Between-group Δ (adj;<br>95% CI; P)* | In Aspirin<br>plus<br>Rivaroxaban,<br>(mean [95%<br>CI]; P)† | In Aspirin<br>Alone,<br>(mean [95%<br>CI]; P)† |
| Total cholesterol, mg/dL |  |  |  |  |  |  |
| Baseline | 136.5±26.0<br>[46] | 137.8±31.0<br>[46] | -1.3 [-13.2,<br>10.6]; P=0.83 | — | — | — |
| 12 months | 145.4±45.9<br>[45] | 138.8±30.9<br>[46] | 6.6 [-9.8,<br>23.0];<br>P=0.42 | — | — | — |
| Absolute<br>change | 8.1±40.4 [45] | 1.0±21.5<br>[46] | 7.1 [-6.5 to<br>20.7];<br>P=0.30 | 7.0 [-6.3,<br>20.3];<br>P=0.30 | 8.1 [-4.0,<br>20.2]; P=0.18 | 1.02 [-5.4,<br>7.4];<br>P=0.75 |
| % change | 6.3±26.6 [45] | 1.9±14.3<br>[46] | 4.4 [-4.6,<br>13.3]<br>P=0.34 | 4.3 [-4.5,<br>13.0];<br>P=0.33 | 6.3 [-1.7,<br>14.3];<br>P=0.12 | 1.9 [-2.3, 6.2];<br>P=0.36 |
| Triglyceride, mg/dL |  |  |  |  |  |  |
| Baseline | 117.5±52.6<br>[46] | 111.3±50.5<br>[46] | 6.2 [-15.2,<br>27.5]; P=0.57 | — | — | — |
| 12 months | 119.5±68.9<br>[45] | 106.7±58.1<br>[45] | 12.8 [-13.9,<br>39.5]; P=0.34 | — | — | — |
| Absolute<br>change | 0.7±47.1 [45] | -5.8±45.7<br>[45] | 6.6 [-12.9,<br>26.0]; P=0.50 | 7.4 [-11.9,<br>26.8]; P=0.45 | 0.7 [-13.4,<br>14.9]; P=0.92 | -5.8 [-19.5,<br>7.9]; P=0.40 |
| % change | 2.1±36.1 [45] | -2.2±34.6 [45] | 4.4 [-10.4, 19.2]; P=0.56 | 5.0 [-9.8, 19.8]; P=0.50 | 2.1 [-8.7, 13.0]; P=0.69 | -2.2 [-12.6, 8.2]; P=0.67 |
| **HDL cholesterol, mg/dL** |  |  |  |  |  |  |
| Baseline | 47.9±12.5 [46] | 50.7±13.5 [46] | -2.7 [-8.1, 2.7]; P=0.32 | — | — | — |
| 12 months | 50.3±10.8 [45] | 52.6±10.7 [45] | -2.2 [-6.8, 2.3]; P=0.33 | — | — | — |
| Absolute change | 2.7±6.9 [45] | 3.1±6.7 [45] | -0.4 [-3.2, 2.5]; P=0.80 | -0.8 [-3.4, 1.8]; P=0.53 | 2.7 [0.6, 4.8]; P=0.01 | 3.1 [1.1, 5.1]; P<0.01 |
| % change | 9.2±25.2 [45] | 7.7±15.0 [45] | 1.5 [-7.2, 10.3]; P=0.72 | -0.1 [-7.7, 7.4]; P=0.97 | 9.2 [1.6, 16.8]; P=0.02 | 7.7 [3.2, 12.2]; P<0.01 |
| **LDL cholesterol, mg/dL** |  |  |  |  |  |  |
| Baseline | 68.8±24.4 [46] | 68.4±27.1 [46] | 0.4 [-10.3, 11.1]; P=0.95 | — | — | — |
| 12 months | 74.9±39.1 [45] | 65.1±20.1 [45] | 9.7 [-3.4, 22.8]; P=0.14 | — | — | — |
| Absolute change | 5.1±36.2 [45] | -2.2±21.0 [45] | 7.3 [-5.2, 19.7]; P=0.25 | 8.3 [-3.3, 20.0]; P=0.16 | 5.1 [-5.8, 16.0]; P=0.35 | -2.2 [-8.5, 4.1]; P=0.48 |
| % change | 10.6±47.7 [45] | 2.5±28.1 [45] | 8.1 [-8.4, 24.5]; P=0.33 | 9.3 [-6.3, 25.0]; P=0.24 | 10.6 [-3.8, 24.9]; P=0.14 | 2.5 [-6.0, 10.9]; P=0.56 |
| **hsCRP, mg/dL** |  |  |  |  |  |  |
| Baseline | 0.10±0.12 [46] | 0.10±0.14 [46] | -0.01 [-0.06, 0.05]; P=0.85 | — | — | — |
| 12 months | 0.09±0.18 [41] | 0.24±0.40 [43] | -0.15 [-0.29, -0.02]; P=0.03 | — | — | — |
| Absolute change | -0.00±0.14 [41] | 0.13±0.42 [43] | -0.14 [-0.28, -0.01]; P=0.04 | -0.15 [-0.28, -0.01]; P=0.03 | -0.01 [-0.05, 0.03]; P=0.66 | 0.13 [0.00, 0.26]; P=0.05 |
| % change | 14.4±104.8<br>[41] | 247.3±585.1<br>[43] | -232.8 [-<br>415.6, -50.1];<br>P=0.01 | -237.6 [-<br>421.5, -53.6];<br>P=0.01 | 14.4 [-18.6,<br>47.5]; P=0.38 | 247.3 [67.2,<br>427.3];<br>P=0.01 |

Data are presented as mean±SD or mean (95% CI). Analysis includes all 92 randomized patients (intention-to-treat population). All analyses were performed on available data without imputation (complete case analysis). Sample sizes [n] indicate the number of patients with available data at each time point.
\*Between-group differences: rivaroxaban plus aspirin group minus aspirin alone group.
Unadjusted differences were assessed using Welch's t-test. Adjusted differences were assessed using ANCOVA with treatment group as the independent variable and baseline value as a covariate. For absolute changes, the model adjusted for baseline measurement; for percent changes, the model adjusted for baseline value.
†Within-group changes assessed using paired t-tests for absolute changes and one-sample t-tests for percent changes.
Abbreviations: HDL, high-density lipoprotein; hsCRP, high-sensitivity C-reactive protein; LDL, low-density lipoprotein; TG, triglycerides

### Safety

No deaths, intracranial hemorrhages, or major bleeding events (BARC 3–5) were observed in either group. Minor bleeding (BARC 1–2) occurred in four patients (8.7%) receiving combination therapy and one patient (2.2%) receiving aspirin alone (P=0.36); all events were epistaxis or gingival bleeding (Table S1).

### Sensitivity and Subgroup Analyses

The per-protocol analysis included 80 patients who completed 12-month ¹⁸F-FDG PET/CT imaging, with mean medication adherence exceeding 90% (92.3% in the combination group; 97.4% in the aspirin-alone group) (Table S2). Results for both primary and secondary endpoints were consistent with the primary analysis (Tables S3–S4): percent change in index carotid MDS TBR was −7.26% (95% CI, −11.04 to −3.38) versus −7.36% (95% CI, −11.06 to −3.67); adjusted difference −1.44% (95% CI, −6.21 to 3.32; P=0.55). Analyses based on absolute TBR changes yielded similar results.

Prespecified subgroup analyses showed no evidence of treatment effect heterogeneity across age, sex, diabetes status, laboratory biomarker, statin intensity, or presence of cardiovascular disease; all treatment-by-subgroup interaction P values were >0.10 (Figure 4). Restricted cubic spline analyses showed no treatment-by-baseline carotid MDS TBR interaction (unadjusted P=0.08; baseline-adjusted P=0.41; Figure S3).

**Figure 4.**
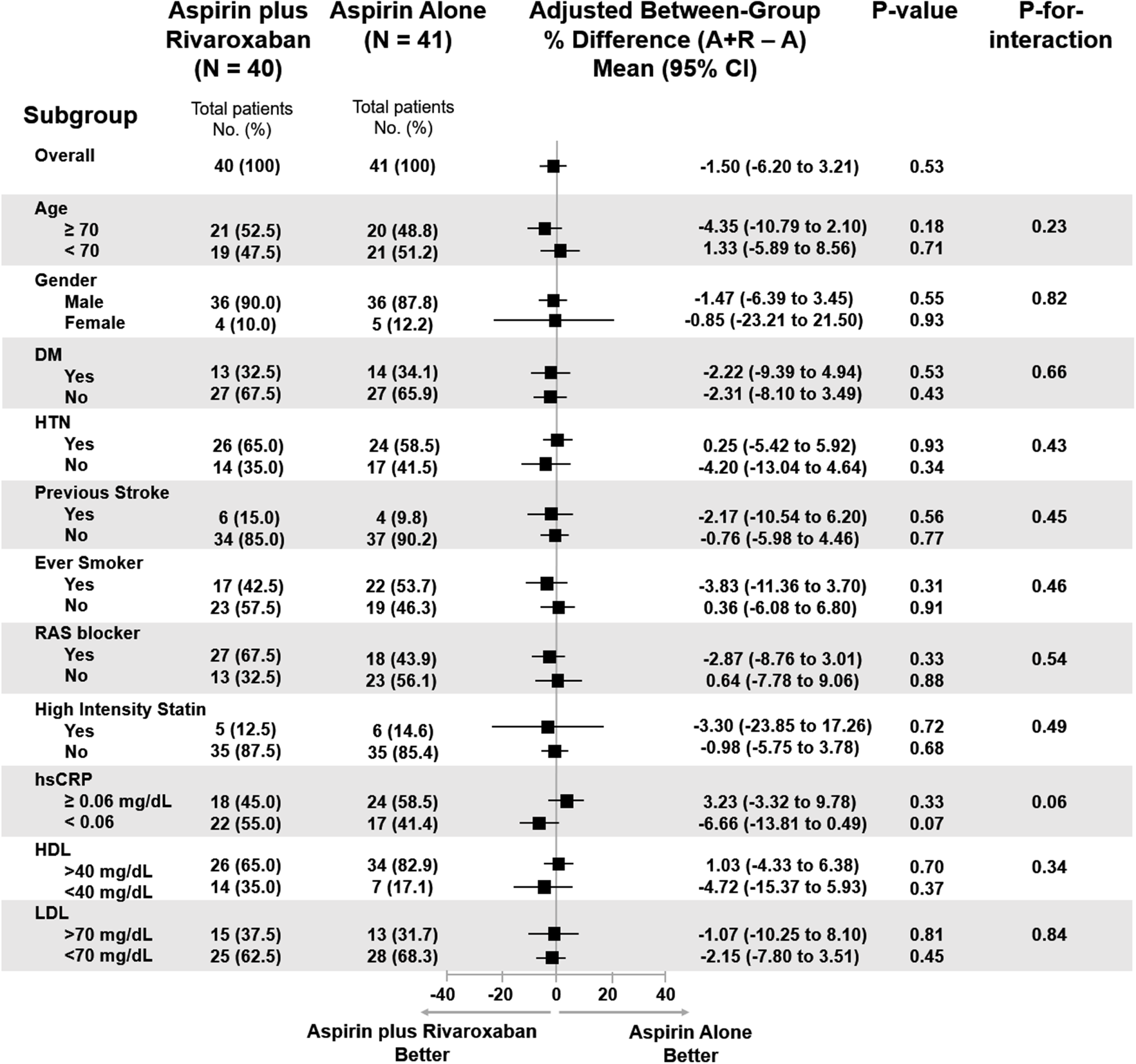
Subgroup Analysis of Treatment Effect on Carotid Artery Inflammation. Percent mean difference (95% CI) in index carotid MDS TBR at 12 months for rivaroxaban plus aspirin versus aspirin alone across prespecified subgroups. Squares represent mean differences; horizontal lines indicate 95% CIs. The vertical line at 0 indicates no treatment difference. Negative values favor rivaroxaban plus aspirin. P-for-interaction values assess heterogeneity of treatment effect across subgroup levels. No significant interactions were observed (all P>0.05). **Abbreviations:** CI, Confidence interval; MDS, most diseased segment; TBR, target-to-background ratio

## DISCUSSION

In this single-center, randomized, open-label trial with blinded imaging endpoints, we evaluated whether adding low-dose rivaroxaban (2.5 mg twice daily) to aspirin attenuates arterial inflammation beyond aspirin alone in patients with asymptomatic carotid atherosclerosis and elevated baseline ¹⁸F-FDG uptake. The primary outcome—percent change in carotid MDS TBR at 12 months—showed no difference between groups (−7.25% vs −7.36%; adjusted between-group difference, −1.50 percentage points; 95% CI, −6.20 to 3.21; P=0.53), with comparable within-group reductions in both arms. Similarly, secondary imaging endpoints, including carotid whole-vessel and ascending aorta TBR, showed no significant between-group differences despite modest within-group reductions. Prespecified subgroup and per-protocol analyses remained consistent with the primary findings, with no evidence of treatment effect heterogeneity across clinical subgroups or the baseline TBR spectrum (P for continuous interaction=0.41). Minor bleeding occurred more frequently with combination therapy (8.7% vs. 2.2%), while no major bleeding events or deaths were observed.

These findings provide mechanistic insights into the cardiovascular benefits observed with dual-pathway inhibition in the COMPASS trial.^18^ Among patients with stable atherosclerotic disease receiving guideline-directed medical therapy, our results suggest that the clinical efficacy of rivaroxaban plus aspirin is mediated primarily through enhanced antithrombotic effects rather than attenuation of macrophage-mediated arterial inflammation at the evaluated dose. Although Factor Xa activates PAR-dependent inflammatory signaling cascades in vascular and inflammatory cell,^19,20^ the anti-inflammatory effects observed in preclinical models did not translate into differential reductions in ^18^F-FDG–assessed plaque metabolism over 12 months in human participants. The similar within-group TBR reductions observed in both treatment arms likely reflect the anti-inflammatory effects of background therapies—particularly statins, which consistently reduce arterial FDG uptake in clinical imaging studies—combined with regression toward the mean inherent in repeated measurements. In addition, aspirin exerts cyclooxygenase-independent anti-inflammatory properties, which may contribute to the observed within-group reductions in both arms.^21^

The consistency of findings across vascular beds strengthens the validity of our neutral primary outcome. Although a modest reduction in hsCRP was observed with combination therapy compared with aspirin alone, this systemic effect did not correspond to differential changes in plaque inflammation, highlighting the dissociation between circulating inflammatory markers and tissue-level arterial inflammation. This dissociation between systemic and local inflammatory markers has been observed in prior studies of lipid-lowering and anti-inflammatory interventions, where changes in circulating biomarkers did not consistently parallel changes in arterial FDG uptake.^22^ Consistent results across prespecified subgroups—including patients with higher baseline inflammation, who may be hypothesized to benefit more—along with the lack of treatment-by-baseline-TBR interaction on a continuous scale (P for interaction=0.41), further support the lack of differential effect of combination therapy on imaging endpoints.

Clinically, among patients with stable atherosclerotic disease receiving guideline-directed medical therapy, the decision to initiate dual-pathway inhibition should be based on assessment of thrombotic risk and bleeding propensity rather than anticipated anti-inflammatory effects on arterial imaging. Patients with elevated ischemic risk—including those with polyvascular disease, peripheral artery disease, or diabetes with end-organ complications—may benefit from dual-pathway inhibition for thrombotic risk reduction.^23,24^ In contrast, patients with inflammatory-predominant phenotypes may respond better to targeted anti-inflammatory interventions.^25,26^ These findings underscore the importance of developing more personalized primary prevention strategies in atherosclerotic cardiovascular disease.

Our findings open several important avenues for future investigation in atherosclerosis treatment. Given that the cardiovascular benefits of dual-pathway inhibition appear mechanistically distinct from anti-inflammatory effects, combining this approach with targeted anti-inflammatory agents (such as colchicine or interleukin-1β antagonists) warrants evaluation. Such combination therapy may provide synergistic cardiovascular protection through complementary mechanisms. To optimize patient selection for such complementary strategies, biomarker-stratified or imaging-enriched trial designs could identify subsets most likely to benefit from adjunctive anti-inflammatory therapy. Furthermore, multimodal imaging approaches that integrate ^18^F-FDG PET with morphological or compositional plaque assessment may better characterize the full spectrum of the vascular effects of dual-pathway inhibition beyond metabolic activity alone. Although higher rivaroxaban doses may theoretically enhance anti-inflammatory effects, the increased bleeding risk precludes such dose escalation in stable disease, highlighting the need for alternative anti-inflammatory strategies.

This study has some limitations. First, as a single-center PROBE trial powered for an imaging surrogate endpoint, our study may lack sufficient power to detect subtle between-group differences or infrequent clinical events. Second, although the centralized blinded image analysis addressed potential bias, the use of a single reader precluded assessment of inter-observer reliability. Third, the 12-month follow-up may have been insufficient to capture delayed anti-inflammatory effects, although prior interventions typically demonstrate FDG changes within 3–6 months. Fourth, external validity is limited by enrollment of predominantly elderly Asian men with moderate carotid stenosis receiving intensive statin therapy. Finally, ¹⁸F-FDG PET/CT predominantly reflects macrophage metabolic activity and may not capture other relevant aspects of plaque pathobiology potentially influenced by Factor Xa inhibition (such as thrombogenicity, neovascularization, and endothelial dysfunction).

In conclusion, among patients with asymptomatic carotid atherosclerosis, the addition of low-dose rivaroxaban to aspirin did not reduce arterial wall inflammation compared with aspirin alone as measured by ¹⁸F-FDG PET/CT over 12 months, suggesting that the cardiovascular benefits observed in prior trials were mediated through antithrombotic rather than anti-inflammatory mechanisms. However, the single-center design, relatively short follow-up period, and focus on imaging endpoints rather than clinical outcomes limit the generalizability of these findings; hence, larger multicenter studies with longer follow-up are needed to fully characterize the mechanistic effects of dual-pathway inhibition in atherosclerosis.

### Authors’ contributions

Conception and design — CW Lee, SW Lee; analysis and interpretation of data — TO Kim, JB Lee; drafting of the manuscript — TO Kim, S Han; critical revision of the manuscript for important intellectual content — CW Lee, JG Gwon, SW Lee; final approval of the manuscript — JG Gwon, SW Lee; statistical expertise — JB Lee; administrative, technical, or logistic support — TO Kim; acquisition of data — TO Kim, S Han, SJ Kang.

## NON-STANDARD ABBREVIATIONS AND ACRONYMS

CI: confidence interval
^18^F-FDG PET/CT: ^18^F-fluorodeoxyglucose positron emission tomography/computed tomography
ANCOVA: Analysis of covariance
HDL: High-density lipoprotein
hsCRP: High-sensitivity C-reactive protein
ITT: Intention-to-treat
LDL: Low-density lipoprotein
MDS: Most diseased segment
PAD: Peripheral artery disease
PAR: Protease-activated receptor
PROBE: Prospective, randomized, open-label, blinded endpoint
SD: Standard deviation
SUV: Standardized uptake value
SVC: Superior vena cava
TBR: Target-to-background ratio

## ACKNOWLEDGEMENTS

We thank the participants, the independent data and safety monitoring board, and the clinical events committee.

## DATA AVAILABILITY STATEMENT

De-identified data and analytic code will be made available upon reasonable request to the corresponding author after publication.

## SUPPLEMENTARY MATERIALS

Supplementary Material Section A–I, Supplementary Tables S1–S4, and Supplementary Figures S1–S3

